# Patterns and correlates of HIV pre-exposure prophylaxis use among higher-risk men who have sex with men in the Jakarta Metropolitan Area, Indonesia

**DOI:** 10.64898/2026.10.12.26364848

**Authors:** Yong Gun Lee, Laura Nevendorff, Wawa Reswana, Tak Ying Wong, Xiaotong Yang, Yue Xu, Chenxi Yang, Yiqiu Huang, Jeffry Oktavianus, Elwin Wu

## Abstract

**Background:** Men who have sex with men (MSM) in the Jakarta Metropolitan Area (JMA), Indonesia are disproportionately affected by HIV. Pre-exposure prophylaxis (PrEP) is a viable strategy for addressing enduring HIV disparities, yet remains underutilized among MSM in the JMA. Patterns and influences of PrEP use may inform future intervention targets but have not been extensively studied in this context.

**Methods:** Guided by the Andersen’s Behavioral Model for Health Services Use, the present study investigated multidimensional correlates of varied PrEP use patterns among 183 JMA-based MSM aged 18-51 (median age=29) and at elevated risk of HIV infection. The multinomial regression model identified factors associated with current PrEP use and some use in the past, compared with never use (reference category).

**Results:** In the present study, 107 (58%) participants reported never use, 34 (19%) current use, and 42 (23%) some use of PrEP. Current PrEP use was negatively associated with HIV stigma and positively with perceived social support, online HIV information seeking, and sexualized drug use. Some PrEP use was negatively associated with HIV stigma and positively with online HIV information seeking.

**Conclusions:** The study findings provide support for the Andersen model’s emphasis on predisposing characteristics, enabling resources, and need factors when tailoring PrEP implementation for higher-risk MSM in the JMA. Addressing stigma, reinforcing social support and HIV knowledge, and integrating PrEP care within multiple health care systems appear viable avenues for improving PrEP adoption in this key population.

## INTRODUCTION

The Jakarta Metropolitan Area (JMA) of Indonesia, covering the capital city of Jakarta and the satellite cities of Bogor, Depok, Tangerang, and Bekasi, has the highest concentrations of MSM in the country and HIV cases among them (Kementerian Kesehatan RI [KEMENKES], 2024). As of 2023, the estimates for HIV prevalence in the MSM populations in the JMA ranged from 15% in Depok to 42% in Tangerang (KEMENKES, 2024). HIV pre-exposure prophylaxis is a highly effective antiretroviral medication (Fonner et al., 2016; Grant et al., 2010) and offers a viable solution to the indicated HIV prevention needs in this context.

Research to date, however, has documented considerable gaps in PrEP use among MSM in Indonesia. PrEP first became available in Indonesia through a pilot implementation program deployed nationally in 2021-2023 for the key populations, including MSM, at elevated risk of HIV infection (Hanum et al., 2025). The implementation program found low levels of uptake and retention among its MSM participants, with more than half of those eligible for PrEP (52%) not initiating it and most of those initiating it (85%) being lost to follow-up within 12 months (Wisaksana et al., 2026). Outside of the implementation program setting, a large online survey of PrEP use among MSM in 15 countries across East, South, and Southeast Asia revealed that the rate of current use in Indonesia, estimated at 8%, was among the lowest in the region (Chan et al., 2026). This is despite prior evidence showing relatively high levels of interest in PrEP use (Bavinton et al., 2021; Cempaka et al., 2020) and thus warrants consideration of other potential factors driving the use in this population.

Research has begun to characterize determinants of PrEP use among higher-risk MSM in Indonesia. Additional analyses of the implementation program outcomes have indicated older age, higher risk perception, and greater PrEP availability as facilitators of initiating the medication among the MSM participants (Wisaksana et al., 2026). Beyond initiation, persistence is equally valuable for achieving the effectiveness of PrEP for HIV prevention (Rutstein et al., 2020; Zhang et al., 2022) but currently remains a challenge among MSM in Indonesia. To illustrate, the implementation program found high rates of seroconversion among the MSM participants with PrEP discontinuation or non-adherence (Hanum et al., 2025), further emphasizing persistence as an unmet need in this population that must be promptly addressed. However, factors contributing to PrEP discontinuation among MSM in Indonesia have not been sufficiently examined.

A wide adoption of PrEP among key populations, including higher-risk MSM, constitutes a principal strategy of HIV prevention in Indonesia (KEMENKES, 2023). Understanding the influence of multidimensional factors in PrEP use is crucial for implementing this strategy and acutely needed in rapidly expanding hotspots for HIV infection such as the JMA. Thus, the present study investigated theoretically relevant factors associated with varied PrEP use patterns, including current use and some use in the past, among higher-risk MSM in the JMA.

## METHODS

### Theoretical Framework

Andersen’s Behavioral Model for Health Services Use emphasizes that the use of health care services is influenced by an array of population and individual characteristics categorized into three dimensions (Andersen, 1995): *predisposing characteristics* refer to demographic characteristics, socio-structural determinants of one’s social status, and health beliefs; *enabling resources* refer to individual and community assets that facilitate health service use; and *need factors* refer to indicators of necessity, as judged by self or others (e.g., medical professionals), of health service use. The Andersen model has provided a framework for understanding the multidimensional factors shaping PrEP use among MSM in the region (Gebregziabher et al., 2026; Mayo & Huang, 2025).

### Study Design, Procedures, and Participants

This is a secondary analysis of data from a study that used a cross-sectional survey design to explore HIV treatment and prevention care service utilization among MSM in the JMA. For the parent study, we applied the following inclusion criteria to determine the study eligibility of survey respondents: being 18 years or older, residing in the JMA, being identified as cisgender man (i.e., self-reporting being assigned male at birth and currently identifying as man), reporting consensual sex with another man within the past 12 months, and providing informed consent.

Participant recruitment and data collection were conducted during April and May 2025 with the support of trained staff that included individuals having direct experience with and access to MSM in the JMA. The staff recruited survey respondents from online and offline venues as well as social networks branded towards MSM, screened those expressing an interest in the study for their study eligibility, and administered a structured survey to those meeting the inclusion criteria.

Participant recruitment yielded contact with 379 individuals initiating the survey; among them, 13 did not meet the inclusion criteria, including two who did not provide informed consent, resulting in a sample of 366 study participants. For the present study, we analyzed the responses of participants who would benefit most from PrEP; this subsample included 183 individuals self-reporting HIV-negative or unknown status and any anal sex encounter with another man within the past three months.

### Ethics

All study procedures and materials were approved by research ethics committees of The University of Hong Kong (EA240385) and Atma Jaya Catholic University of Indonesia. Informed consent was obtained from all study-eligible participants at the start of the survey. Monetary compensation of 50,000 Indonesian Rupiah (IDR) was provided to participants upon completion of the survey.

### Measures

Participants were asked whether they had ever, in lifetime, used PrEP. Response options included “never”, “yes, I am currently taking it” (i.e., current use), and “yes, I took it in the past but stopped” (i.e., some use).

Measures of predisposing characteristics included age, relationship status, education level, employment status, and HIV stigma. <u>Age</u> was self-reported in years and ranged between 18 and 51 (Median=29). <u>Relationship status</u> was assessed with a single item with the following response options: “single, never married”, “in a relationship (not legally married)”, “married”, “divorced”, “widowed”, “separated”, and “other, specify”. <u>Education level</u> was operationalized as the highest level of formal education completed and assessed with an item with the following response options: “did not complete senior high school”, “completed senior high school”, “completed undergraduate school”, “completed graduate school”, and “other, specify”. <u>Employment status</u> was assessed with an item with the following response options: “unemployed, looking for work”, “unemployed, not looking for work”, “temporary leave from work”, “employed or self-employed, full-time”, “employed or self-employed, part-time”, “retired”, “student”, and “other, specify”. <u>HIV stigma</u> was assessed using the 6-item Attitude Towards People Living with HIV (PLHIV) scale (Nyblade et al., 2013). The items, including “Most people living with HIV do not care if they infect other people” and “People living with HIV should feel ashamed of themselves,” were scored on a 4-point Likert scale ranging from “strongly disagree” (0) to “strongly agree” (3). Scores across the items were summed to obtain a total score measuring HIV stigma (α=.79); higher total scores indicated greater stigmatizing attitude towards people living with HIV.

Measures of enabling resources included city of residence, income level, health insurance coverage, perceived social support, and online HIV information seeking. <u>City of residence</u> was self-reported as currently living in Jakarta, Bogor, Depok, Tangerang, or Bekasi. <u>Income level</u> was operationalized as average monthly income during the past six months and assessed with an item with following response options: “less than 2.5 million IDR per month”, “2.5 million – 5 million IDR per month”, and “more than 5 million IDR per month”. <u>Health insurance coverage</u> was assessed with a binary item indicating having or not having health insurance coverage. <u>Perceived social support</u> was assessed using the 6-item short version of the Multidimensional Scale of Perceived Social Support (MSPSS) scale (Wu et al., 2025; Zimet et al., 1990). The items, including “There is a special person who is around when I am in need” and “My friends really try to help me,” were scored on a 7-point Likert scale ranging from “very strongly disagree” (0) to “very strongly agree” (6). Scores across the items were summed to obtain a total score measuring perceived social support (α=.75); higher total scores indicated greater levels of perceived social support. <u>Online HIV information seeking</u> was assessed with a binary item indicating having or not having sought HIV-related information on the internet within the past two weeks.

Measures of need factors included HIV-risk-associated sex behaviors. <u>Sexually transmitted infection (STI) diagnosis</u> was assessed with a binary item indicating being or not being diagnosed with an STI, other than HIV, within the past six months. Four additional <u>behavioral indicators of HIV risk</u> were assessed using items adapted from the U.S. National Institute on Drug Abuse HIV Risk Behaviors questionnaire (National Institute on Drug Abuse [NIDA], 2013). First, participants self-reported the number of men they had sex with within the past three months and, if reporting at least one sex partner, the total number of anal sex encounters with them. Participants reporting at least once anal sex encounter were further probed about the frequencies of condom use, sexualized drug use, and transactional sex; for these items, response options included “never”, “less than half of the time”, “more than half of the time”, and “every time”.

### Study Variables

Participant responses for the “other, specify” category in the measures of relationship status, education level, and employment status were reviewed and, as applicable, inductively coded by authors (YGL, LN, WR) as predefined response options. Responses for most study variables were collapsed to enhance interpretation. Relationship status was dichotomized as being in a partnered relationship (1) if reporting being in a relationship or married and not being in a partnered relationship (0) if reporting being single, divorced, widowed, or separated. Education level was dichotomized as completing undergraduate or higher-level education (1) and high school or lower-level education (0). Employment status was dichotomized as being currently employed (1) and not currently employed (0) if reporting being unemployed, on temporary leave, retired, or a student. Income was dichotomized as earning minimum wage (1) if reporting monthly income of more than IDR 5 million and less than minimum wage (0) if reporting monthly income of IDR 5 million or less. The number of male sex partners was dichotomized as having a singular sex partner (0) if reporting having sex with one person and having multiple sex partners (1) if reporting having sex with two or more people within the past three months. Condom use frequency was dichotomized as using a condom every time (0) and any condomless anal sex (1) if reporting condom use at a lesser frequency, including never, less than half of the time, and more than half of the time. Frequencies of sexualized drug use and transactional sex were each dichotomized as never (0) and any engagement (1). Age and the total scores of the Attitude Towards PLHIV scale and the short version MSPSS scale were treated as continuous variables.

### Data Analysis

Descriptive statistics were used to summarize participants’ PrEP use patterns (never use, current use, some use) as well as sociodemographic, psychosocial, and behavioral characteristics across the Andersen model domains of predisposing characteristics, enabling resources, or need factors. Differences in PrEP use patterns by Andersen model factors were evaluated using chi-square tests for the categorical variables and one-way ANOVA for the continuous variables. A multivariable multinomial logistic regression model was used to examine the associations between Andersen model factors and current use and some use of PrEP, with never use as the reference category. Adjusted odds ratios (AORs) and their 95% confidence intervals (CIs) were estimated, with statistical significance set at *p*<.05. All statistical analyses were performed using SPSS version 29.0.

## RESULTS

The results of the univariate and bivariate analyses are presented in Table 1. Participants came from all five cities, with the highest proportion residing in Jakarta (*n*=45; 25%) and the lowest proportion in Tangerang (*n*=28; 15%). Participant ages ranged from 18 to 51 years, with the median age of 29 years. Lower proportions of the study participants were in a partnered relationship (*n*=32; 17.5%), had completed university-level education (*n*=37; 20%), and had engaged in online HIV information seeking within the past two weeks (*n*=69; 38%). Higher proportions were employed (*n*=123; 67%), earned minimum wage or less over the past six months (*n*=132; 72%), and had health insurance coverage (*n*=146; 80%).

**Table 1.** Study sample characteristics, full sample and by PrEP use patterns (*N*=183)

|  | Full sample<br>*M (SD) / n (%)* | Never use<br>(*n*=107; 58.5%) | Current use<br>(*n*=34; 18.6%) | Some use<br>(*n*=42; 23.0%) |
| --- | --- | --- | --- | --- |
| **Predisposing characteristics** |  |  |  |  |
| Age, in years (Range=18-51, Median=29) | 31.1 (8.3) | 30.2 (8.4) | 32.1 (8.0) | 32.6 (8.4) |
| Relationship status |  |  |  |  |
| Single, divorced, widowed, separated | 151 (82.5%) | 89 (83.2%) | 28 (82.4%) | 34 (81.0%) |
| Married, in a relationship | 32 (17.5%) | 18 (16.8%) | 6 (17.6%) | 8 (19.0%) |
| Education status |  |  |  |  |
| High school or lower completed | 146 (79.8%) | 90 (84.1%) | 27 (79.4%) | 29 (69.0%) |
| University or higher completed | 37 (20.2%) | 17 (15.9%) | 7 (20.6%) | 13 (31.0%) |
| Employment status |  |  |  |  |
| Unemployed, temporary leave, retired, student | 60 (32.8%) | 41 (38.4%) | 10 (29.4%) | 9 (21.4%) |
| Employed | 123 (67.2%) | 66 (61.7%) | 24 (70.6%) | 33 (78.6%) |
| HIV stigma ($\alpha=.79$) | 9.2 (3.3) | 9.6 (3.4) | 8.6 (3.4) | 8.7 (3.1) |
| **Enabling resources** |  |  |  |  |
| City of residence |  |  |  |  |
| Jakarta | 45 (24.6%) | 29 (27.1%) | 7 (20.6%) | 9 (21.4%) |
| Bogor | 36 (19.7%) | 25 (23.4%) | 5 (14.7%) | 6 (14.3%) |
| Depok | 43 (23.5%) | 21 (19.6%) | 9 (26.5%) | 13 (31.0%) |
| Tangerang | 28 (15.3%) | 15 (14.0%) | 4 (11.8%) | 9 (21.4%) |
| Bekasi | 31 (16.9%) | 17 (15.9%) | 9 (26.5%) | 5 (11.9%) |
| Income, past 6 months |  |  |  |  |
| Minimum wage (IDR 5 million) or less | 132 (72.1%) | 82 (76.6%) | 24 (70.6%) | 26 (61.9%) |
| Higher than minimum wage | 51 (27.9%) | 25 (23.4%) | 10 (29.4%) | 16 (38.1%) |
| Health insurance coverage |  |  |  |  |
| No | 37 (20.2%) | **29 (27.1%)** | **6 (17.6%)** | **2 (4.8%)** |
| Yes | 146 (79.8%) | **78 (72.9%)** | **28 (82.4%)** | **40 (95.2%)** |
| Perceived social support ($\alpha=.75$) | 13.0 (4.6) | **12.7 (4.3)** | **14.4 (4.9)** | **12.8 (5.0)** |
| Online HIV information seeking, past 2 weeks |  |  |  |  |
| No | 114 (62.3%) | **78 (72.9%)** | **17 (50.0%)** | **19 (45.2%)** |
| Yes | 69 (37.7%) | **29 (27.1%)** | **17 (50.0%)** | **23 (54.8%)** |
| **Need factors** |  |  |  |  |
| STI diagnosis, past 6 months |  |  |  |  |
| No | 156 (85.2%) | 96 (89.7%) | 27 (79.4%) | 33 (78.6%) |
| Yes | 27 (14.8%) | 11 (10.3%) | 7 (20.6%) | 9 (21.4%) |
| Multiple sex partnership, past 3 months |  |  |  |  |
| No | 48 (26.2%) | 27 (25.2%) | 9 (26.5%) | 12 (28.6%) |
| Yes | 135 (73.8%) | 80 (74.8%) | 25 (73.5%) | 30 (71.4%) |
| Condomless anal sex, past 3 months |  |  |  |  |
| No | 60 (32.8%) | 30 (28.0%) | 12 (35.3%) | 18 (42.9%) |
| Yes | 123 (67.2%) | 77 (72.0%) | 22 (64.7%) | 24 (57.1%) |
| Sexualized drug use, past 3 months |  |  |  |  |
| No | 136 (74.3%) | **82 (76.6%)** | **19 (55.9%)** | **35 (83.3%)** |
| Yes | 47 (25.7%) | **25 (23.4%)** | **15 (44.1%)** | **7 (16.7%)** |
| Transactional sex, past 3 months |  |  |  |  |
| No | 136 (74.3%) | 76 (71.0%) | 26 (76.5%) | 34 (81.0%) |
| Yes | 47 (25.7%) | 31 (29.0%) | 8 (23.5%) | 8 (19.0%) |
| | | | | Bold denotes $p < .05$ |

Within the past three months, higher proportions of the study participants had multiple sex partners (*n*=135; 74%) and had ever engaged in condomless anal sex (*n*=123; 67%), and lower proportions had ever engaged in sexualized drug use (*n*=47; 26%) and transactional sex (*n*=47; 26%). Fewer participants had a sexually transmitted infection diagnosis within the past six months (*n*=27; 15%).

Regarding PrEP use, 107 (58%) participants reported never use, 34 (19%) current use, and 42 (23%) some use. The PrEP use patterns varied statistically significantly by several factors across the Andersen model domains, including health insurance coverage, perceived social support, online HIV information seeking, and sexualized drug use. Health insurance coverage was the lowest among the participants reporting never use and the highest among those reporting some use (X^2^=9.5, *p*<.01). The mean score for the short-version MSPSS scale was the lowest among the participants reporting never use and the highest among those reporting current use (*F*(2,180)=3.6, *p*=.03). The rate of online HIV information seeking within the past two weeks was the lowest among the participants reporting never use and the highest among those reporting some use (X^2^=12.5, *p*<.01). Finally, the rate of sexualized drug use within the past three months was the lowest among the participants reporting some use and the highest among those reporting current use (X^2^=8.1, *p*=.02).

The results from the multinomial regression model are presented in Table 2. Current use of PrEP, compared with never use, was statistically significantly associated with several factors across the Andersen model domains. Among the predisposing characteristics assessed, HIV stigma was associated with lower odds of current use (*AOR*=0.83; *95% CI*=0.69, 0.99). Among the enabling resources, both perceived social support (*AOR*=1.11; *95% CI*=1.01, 1.22) and online HIV information seeking (*AOR*=3.31; *95% CI*=1.30, 8.47) were associated with higher odds of current use. Among the need factors, sexualized drug use was associated with higher odds of current use (*AOR*=4.50; *95% CI*=1.52, 13.27).

**Table 2.** Multinomial regression model examining the associations of current use and some use of PrEP with Andersen model factors among a sample of higher-risk MSM in the JMA (*N*=183)

|  | Current use<br>AOR (95% CI) | Some use<br>AOR (95% CI) |
| --- | --- | --- |
| **Predisposing characteristics** |  |  |
| Age, in years | 1.03 (0.97, 1.09) | 1.01 (0.96, 1.07) |
| Relationship status: Married, in a relationship | 1.06 (0.27, 4.09) | 1.51 (0.43, 5.29) |
| Education status: University or higher completed | 0.91 (0.28, 2.98) | 1.27 (0.45, 3.59) |
| Employment status: Employed | 1.28 (0.46, 3.60) | 1.46 (0.54, 3.94) |
| HIV stigma | **0.83 (0.69, 0.99)** | **0.83 (0.70, 0.98)** |
| **Enabling resources** |  |  |
| City of residence |  |  |
| Jakarta | Ref. | Ref. |
| Bogor | 0.41 (0.07, 2.41) | 0.24 (0.05, 1.19) |
| Depok | 2.33 (0.57, 9.53) | 3.12 (0.87, 11.20) |
| Tangerang | 1.23 (0.23, 6.69) | 1.81 (0.45, 7.30) |
| Bekasi | 2.76 (0.71, 10.76) | 0.80 (0.18, 3.58) |
| Income, past 6 months: Higher than minimum wage | 1.00 (0.31, 3.25) | 1.84 (0.60, 5.66) |
| Health insurance coverage: Yes | 1.31 (0.38, 4.58) | 4.59 (0.88, 23.86) |
| Perceived social support | **1.11 (1.01, 1.22)** | 1.03 (0.94, 1.12) |
| Online HIV information seeking, past 2 weeks: Yes | **3.31 (1.30, 8.47)** | **3.58 (1.49, 8.63)** |
| **Need factors** |  |  |
| STI diagnosis, past 6 months: Yes | 2.55 (0.77, 8.41) | 2.27 (0.71, 7.27) |
| Multiple sex partnership, past 3 months: Yes | 1.61 (0.50, 5.17) | 2.06 (0.67, 6.39) |
| Condomless anal sex, past 3 months: Yes | 0.63 (0.23, 1.71) | 0.48 (0.18, 1.25) |
| Sexualized drug use, past 3 months: Yes | **4.50 (1.52, 13.27)** | 0.72 (0.23, 2.26) |
| Transactional sex, past 3 months: Yes | 0.55 (0.17, 1.82) | 1.24 (0.39, 3.93) |

Bold denotes $p < .05$

Some use of PrEP, compared with never use, was statistically significantly associated with a predisposing characteristic and an enabling resource. Specifically, HIV stigma was associated with lower odds (*AOR*=0.83; *95% CI*=0.70, 0.98) while online HIV information seeking was associated with higher odds (*AOR*=3.58; *95% CI*=1.49, 8.63) of some use.

## DISCUSSION

The proportion of MSM in the present study reporting any prior use of PrEP, including current use and some use in the past, was higher than that observed in previous studies (Chan et al., 2026; Wirawan et al., 2025); the majority, however, were unPrEPed, asserting the value of ongoing efforts to identify potential facilitators and barriers. Guided by the Andersen model, the present study has elucidated the influence of predisposing characteristics, enabling resources, and need factors in varied PrEP use patterns among MSM in the JMA. Specifically, lower HIV stigma, greater perceived social support, online HIV information seeking, and sexualized drug use emerged as significant correlates of current use, compared with never use. Similarly, lower HIV stigma and online HIV information seeking significantly predicted some use in the past, compared with never use.

Findings of the present study should be interpreted with consideration of several methodological limitations. First, the cross-sectional design has limited causal inferences for the associations between the study variables. Next, despite efforts to ensure accuracy in data collection, the self-reporting method could have introduced recall and social desirability biases. Moreover, the assessment of PrEP use patterns as three distinct categories could have masked important nuances. For instance, the experiences and perceptions of PrEP use may differ for individuals reinitiating the medication and by varying levels of adherence and persistence.

Finally, convenience and snowball sampling methods likely compromised the generalizability of the study findings to larger populations of MSM in the JMA. These methods, however, facilitated the inclusion of a diverse sample of MSM in the present study and are generally considered suitable, if not necessary, for recruitment within the contexts of high stigma and mistrust (Ana Gama et al., 2017; Ellard-Gray et al., 2015).

The present study found that greater HIV stigma was negatively linked to current and some PrEP uses, compared with never use. This is consistent with the extant research illustrating that stigma predisposes MSM to PrEP avoidance, thus serving as a significant barrier (Dhir et al., 2026; Elopre et al., 2018). Negative attitude towards PLHIV among MSM without HIV likely reflects misconceptions about HIV (Sjöland et al., 2026) and has been postulated to impede the use of preventive services through engendering fear of being labeled and judged (Boudewyns et al., 2026; Brown et al., 2016). The present study findings could therefore be understood as resulting from PrEP avoidance among individuals anticipating greater HIV-related social risk.

Alternatively, MSM reporting current or some PrEP use could represent individuals equipped with resources to effectively address HIV stigma. A recent study exploring the impact of PrEP initiation among MSM in Singapore demonstrates this possibility, highlighting the contributions of social support and HIV prevention knowledge, garnered through peer interactions, to the significant reductions in stigma related to HIV and PrEP over time (Naidu et al., 2026).

Given the dynamics of stigma, social support, HIV prevention knowledge, and PrEP use, it is not surprising that perceived social support and online HIV information seeking were positively linked to any prior PrEP use. Although we did not directly assess HIV prevention knowledge, it is reasonable to posit that online HIV information seeking led to its acquisition (Veinot et al., 2013). In addition, there is evidence illustrating that online HIV information seeking amplifies social support for MSM to help address their perceived risk of infection and the social risk surrounding HIV (Chen et al., 2019; Threats, 2025). It is possible that MSM reporting current PrEP use in the present study represent individuals continuously involved in the exchange of knowledge and support regarding the mitigation of HIV risk and engagement in the novel antiretroviral medication. The present study findings thus align with the extant research indicating social support and HIV prevention knowledge, including PrEP knowledge, as enabling resources facilitating PrEP use for diverse MSM (Gallardo-Cartagena et al., 2026; Mayo & Huang, 2025; Owens et al., 2022; Sohail et al., 2025). They further underscore the viability of online information seeking for enabling PrEP use among MSM in the JMA, likely through enhancing knowledge and support related to HIV, PrEP, and other stigmatized topics (Nevendorff et al., 2025). Future research should examine in greater detail the types and sources of support and knowledge related to HIV and PrEP.

The co-occurrence of PrEP use with sexualized drug use among MSM in the region is well established in research (Eger et al., 2022; Mayo & Huang, 2025; Sun et al., 2024; Wang et al., 2020; Wirawan et al., 2025; Wong et al., 2025). In line with this, the present study found that sexualized drug use was positively associated with current PrEP use, likely suggesting that MSM adjust their behaviors according to varying perceptions of HIV risk. Research proposes that, in risk reduction scenarios, individuals engaging in sexualized drug use might perceive increased risk of HIV, leading them to adopt PrEP as another layer of protection (Alohan et al., 2024; Gafos et al., 2019; Sohail et al., 2025); this is consistent with the conceptualization of increased risk perception as a need factor driving PrEP use. However, in risk compensation scenarios, individuals using PrEP might expect and achieve decreased risk of HIV, leading them to engage in behaviors that, without adequate protection, would confer risk (Normallairy et al., 2026) but, with protection, safety, pleasure, and other benefits (Murphy et al., 2024; Philbin et al., 2016).

## CONCLUSIONS

The present study findings offer insights into the modifiable targets in future interventions for promoting PrEP adoption among higher-risk MSM in the JMA. Renewed efforts of addressing intersectional stigma in community and clinical settings are warranted, particularly through reframing negative attitudes towards HIV and affirming PrEP use as a health priority and not an indication of deviance. Fostering a culture of knowledge sharing and social support appears essential and should leverage digital information and communication technology used by MSM. In anticipation of multifaceted risk, PrEP care should integrate counseling focused on risk assessment and communication and be integrated within medical care and drug use treatment programs.

## Author Contributions

YGL: Conceptualization, Methodology, Formal analysis, Investigation, Data curation, Writing – Original draft, Supervision, Project administration, Funding acquisition

LN: Investigation, Resources, Writing – Review & editing, Supervision, Project administration

WR: Investigation, Resources, Writing – Review & editing, Project administration

TYW: Resources, Writing – Review & editing XY: Writing – Review & editing

YX: Writing – Review & editing CY: Writing – Review & editing

YH: Writing – Review & editing

JO: Writing – Review & editing

EW: Writing – Review & editing, Supervision

## Acknowledgements

The authors thank the project staff (Aditya, Catur, Dika, Marsal), community consultants, and study participants for their contributions to the study.

## Funding

This study was supported by the Hong Kong Research Grants Council [Early Career Scheme, Project no. 27616725 (PI: Lee)].

## Conflict of Interest

The author(s) declare no potential conflicts of interest.

## Data Availability

Data supporting the conclusions of this study may be made available upon reasonable request.

## Notes

### Competing Interest Statement

The authors have declared no competing interest.

### Author Declarations

Ethics committees of The University of Hong Kong and Atma Jaya Catholic University of Indonesia gave *ethical* approval for this work.

